# *PDGRFA*-mutant gastrointestinal stromal tumours (GISTs) in Eastern England: clinicopathological features and outcomes of 50 patients diagnosed between 2008-2021

**DOI:** 10.1101/2021.12.24.21268386

**Authors:** David M Favara, Han Wong, Jennifer Harrington, Olivier Giger, V Ramesh Bulusu

## Abstract

**Background:** *PDGRFA*-mutant gastrointestinal tumours (GISTs) comprise approximately 10% of GISTs and are mostly gastric. Targeted therapies against these tumours have historically been limited by tyrosine kinase inhibitor (TKI)-resistance. We reviewed the characteristics and outcome of all *PDGRFA*-mutant patients seen at our centre over the last 13 years.

**Methods:** All *PDGRFA*-mutant patients seen at the Cambridge University Hospitals NHS Foundation Trust GIST clinic from July 2008-July 2021 were retrospectively reviewed and followed up.

**Results:** 50 *PDGRFA*-mutant GIST patients were diagnosed during the 13-year period. 60% were male. Median tumour size was 5 cm and the majority were epithelioid (44%) or mixed (36%) type. Commonest primary location was the gastric body (52%). In non-metastatic patients the low-risk modified AFIP risk group was the most common (65.2%). *PDGFRA* exon 18 (86%) were the most common *PDGFRA* mutations, most being imatinib resistant. None harboured a *KIT* mutation. 38% of cases had high KIT expression (immunohistochemistry), whilst 48% had patchy expression and 14% were negative. Most were positive for DOG1 (94%). CD34 was highly expressed in 48% of cases. 13% developed metastases during the 13-year follow-up period: liver (80%) being the commonest site. 76% of all patients underwent radical resection. 14% received TKI therapy. After a median follow-up of 55.1 months, 82% remained alive: 6 patients died from metastatic GIST; 3 from other causes. Median time to metastatic disease was 30.1 months, and median time from metastatic diagnosis to death was 18.5 months. Patients who presented with metastatic disease had the poorest survival outcome (*p*=0.001)

**Conclusion:** To date this is the largest single-centre European *PDGRFA*-mutant GIST cohort. Results highlight the generally indolent nature of *PDGRFA*-mutant disease (contrasted to the poor outcome of those who present with metastatic disease), male and gastric preponderance (unlike *KIT*-mutant GISTs), and variable KIT expression.

**Highlights:**

1. Largest single-centre European *PDGFRA*-mutant GIST series (n=50) to date
2. Results confirm the relative indolent nature of non-metastatic *PDGFRA-mutant* GISTs
3. *PDGFRA*-mutant GISTs had variable levels of KIT protein expression (immunohistochemistry) despite being *KIT*-wild type.
4. In contrast to *KIT*-mutant GISTs, *PDGFRA*-mutant GISTs were more common in males and less frequent in parts of the stomach where *KIT*-mutant GISTS occur.
5. A minority of patients received TKI therapy. One patient with metastatic imatinib-resistant *PDGFRA*-mutant GIST had a 6 year response to imatinib.

## Introduction

Gastrointestinal stromal tumours (GISTs) arise from the interstitial cells of Cajal^1^ and have an annual incidence of between 10-15 cases per million population^2^. The majority (85%) are driven by mutually exclusive mutations in receptor tyrosine kinases (RTKs):^3 4^ KIT (75%) and platelet derived growth factor receptor alpha (*PDGFRA*) (10%). The remaining 15% being caused by non-TKI drivers including *BRAF* and *SDH* mutations^5^.

Almost all *PDGFRA*-mutant GISTs originate in the stomach^6^, compared to 60% of *KIT*-mutant GISTs^7^. *PDGFRA*-mutant GISTs are driven (in order of decreasing frequency) by mutations affecting the following *PDGFRA* regions: exon 18 (activation loop region), exon 12 (juxtamembrane domain) and exon 14 (ATP-binding domain) ^8^. The treatment of PDGFRA-mutant GISTs has historically been limited by inherent tyrosine kinase inhibitor (TKI) - resistance which contrasts to *KIT*-mutant GISTs, where the majority are sensitive to first- and second-generation TKIs. This is, however, changing with the development of new agents. Recently, avapritinib^9^, a third-generation TKI has been shown to have potent effect against exon 18 D842V *PDGFRA*-mutant GISTs (the commonest *PDGFRA* mutation type), was approved by the FDA and EMA for exon 18 D842V *PDGFRA*-mutant GISTs^10^. More recently, ripretinib, a novel TKI switch-pocket inhibitor has been shown to have potent and broad activity against a range of *KIT* and *PDGFRA* mutations^11^, leading to it being licenced as fourth line therapy^12^.

In this study, we present a 13-year retrospective follow-up of 50 newly diagnosed *PDGRFA*-mutant patients seen at a single centre in the United Kingdom (UK). To date this is the largest single-centre European cohort published.

## Methods

### Data collection

Retrospective data collection was performed on all patients who were reviewed by the GIST clinic, Addenbrooke’s Hospital, Cambridge University Hospitals NHS Foundation Trust (CUH) (Cambridge, UK) between July 2008 and July 2021. Data collected included all data relating to diagnosis (included results from mutational-profiling and immunohistochemistry), treatment, and follow-up. Ethical approval was received from the Institutional Review Board.

### Sample collection and immunohistochemistry

Patients underwent either surgical resection or fine-needle aspirate (FNA) under endoscopic ultrasound at the time of their diagnosis. Tissue specimens were formalin-fixed and underwent routine immunological staining for KIT(also known as CD117), DOG1, desmin, S100 and CD34 expression using standardised methods in use in the UK’s National Health Service (NHS). Immunohistochemical results were classified into three expression categories: i) positive (strong positivity in > 90% of tumour cells), ii) patchy (weaker, non-uniform staining < 90% of tumour cells and iii) negative expression. GIST mitoses were counted per 50 high-power fields (HPFs). Risk prediction was evaluated according to the modified Armed Forces Institute of Pathology (AFIP) criteria^13 14^.

### Mutational analyses

Tumour mutation analysis was performed on DNA extracted from FFPE slides as previously described ^15^. All GIST samples underwent routine *KIT* exons 9, 11, 13 and 17 and *PDGFRA* exons 12, 14 and 18 mutational profiling by dideoxy sequencing or NGS panel sequencing using the Ion AmpliSeq™ Cancer Hotspot Panel v2 (ThermoFisher) as per standardised methods as previously described^16^.

### Statistical analyses

Survival analyses were performed in R (version 4.0.3) using the Cox Proportional-Hazards Model.

## Results

Between July 2008 and July 2021 (13 years), 354 histologically-proven individual GIST patients were reviewed at the CUH GIST clinic. Of these, 50 were *PDGRFA*-mutant (50/354; 14.1%). Their characteristics are listed in **Table 1**. The majority of *PDGRFA*-mutant GIST patients were male (60%) and above the age of 50 (88%). Most (76%) were diagnosed at regional hospitals and referred to CUH for histology review, molecular profiling and care. GI bleeding was the commonest presenting complaint (15/50; 30%) followed by anaemia (12/50; 24%). Eight patients (16%) had another type of cancer at the time of the GIST diagnosis or during their follow-up: melanoma (2 patients), lung cancer (2 patients), prostate cancer (1 patient), diffuse large B-cell lymphoma (1 patient), chronic myeloid leukaemia (1 patient), and penile squamous cell carcinoma (1 patient). As part of diagnosis, all patients underwent CT staging and 20% (10/50) also received a PET-CT scan. Most (38/50; 76%) were treated with radical resection. The remainder (12/50; 24%) underwent biopsy without resection due to a number of factors: small volume GIST, poor anaesthesia risk, or the presence of metastatic disease.

**Table 1:** patient demographics

|  | n=50 | % |
| --- | --- | --- |
| **Gender** |  |  |
| Male | 30 | 60.0% |
| Female | 20 | 40.0% |
| **Median age (range)** |  |  |
| Age <50 | 6 (19-48) | 12.0% |
| Age >50 | 44 (50-87) | 88.0% |
| **Referral to centre** |  |  |
| Yes | 38 | 76.0% |
| No | 12 | 24.0% |
| **Presenting problem** |  |  |
| GI bleed | 15 | 30.0% |
| Anaemia | 12 | 24.0% |
| Incidental | 8 | 16.0% |
| Abdominal pain | 9 | 18.0% |
| Unknown | 6 | 12.0% |
| **Other cancers (syn- and metachronous)** |  |  |
| yes | 8 | 16.0% |
| no | 42 | 84.0% |
| **Diagnostic staging** |  |  |
| CT | 50 | 100.0% |
| PET-CT | 10 | 20.0% |
| **Procedure** |  |  |
| Radical resection: laparoscopic | 23 | 46.0% |
| Radical resection: open | 15 | 30.0% |
| Biopsy only | 12 | 24.0% |

**Table 2** lists their tumour and treatment characteristics. Most (49/50; 98%) primary *PDGRFA*-mutant GISTs were gastric with the highest incidence in the gastric body (26/50; 52%) followed by the gastric antrum (11/50; 22%) (**Figure 1**). In 20% of patients (10/50), the exact intra-gastric location of the gastric GIST was not recorded. In one patient who presented with metastatic peritoneal disease, a primary tumour location could not be identified endoscopically or radiologically. Most tumours were smaller than 5 cm (22/50; 44%) and showed an epithelioid histological growth pattern (22/50; 44%). The majority of the 46 patients who presented with limited disease fell into the low risk AFIP group for tumour progression (30/46; 65.2%). Exon 18 mutations were the commonest *PDGFRA* mutations (43/50; 86%), with most being imatinib resistant. None of the *PDGFRA*-mutated GISTs harboured a *KIT* mutation. Immunohistochemistry (IHC) (**supplementary table 1**) showed that 38% of cases were highly KIT(CD117) positive, whilst 48% had patchy KIT expression and 14% were negative. DOG1 was positive in 96% of tumours (47/49; 1 case was negative and 1 case was unrecorded). CD34 was highly expressed in 48% (24/50) of cases.

**Table 2:** Tumour and treatment characteristics

|  | n=50 | % |  | n=50 | % |
| --- | --- | --- | --- | --- | --- |
| **GIST location** |  |  | **Metastatic at diagnosis** |  |  |
| Gastric | 49 | 98.0% | No | 46 | 92.0% |
| Extra-gastric | 1 | 2.0% | Yes | 4 | 8.0% |
| **Gastric location:** |  |  | **Developed metastases during follow-up** |  |  |
| Cardia | 0 | 0.0% | No | 40 | 87.0% |
| Fundus | 1 | 2.00% | Yes | 6 | 13.0% |
| Body | 26 | 52.0% | **Metastatic sites: (n=10 patients)** |  |  |
| Antrum | 11 | 22.0% | Liver | 8 | 80.0% |
| Pylorus | 1 | 2.0% | Peritoneum | 5 | 50.0% |
| Not recorded | 10 | 20.0% | Lung | 1 | 10.0% |
| No GI primary | 1 | 2.0% | Pancreas | 1 | 10.0% |
| **Size:** |  |  | **TKI therapy (n=7 individuals)** |  |  |
| Median cm (mean) | 5 (7.9) | - | Neoadjuvant |  |  |
| <5 | 22 | 44.0% | Imatinib | 1 | 2.2% |
| 5-10 | 14 | 28.0% | **Adjuvant** |  |  |
| >10 | 14 | 28.0% | Adjuvant imatinib | 2 | 4.3% |
| **Mitotic count** |  |  | No treatment | 40 | 87.0% |
| <=5 counts | 40 | 80.0% | **Metastatic** |  |  |
| 5-10 counts | 4 | 8.0% | Imatinib/nilotinib/sunitinib | 3 | 30.0% |
| >10 counts | 6 | 12.0% | Avapritinib | 2 | 20.0% |
| **Modified AFIP risk stratification** |  |  | No treatment | 5 | 50.0% |
| Low risk | 30 | 65.2% | **Survival** |  |  |
| Intermediate risk | 8 | 17.4% | Alive | 41 | 82.0% |
| High risk | 8 | 17.4% | Dead | 9 | 18.0% |
| **Histology** |  |  | **Cause of death** |  |  |
| Epithelioid | 22 | 44.0% | Death GIST cause | 6 | 66.7% |
| Spindle | 10 | 20.0% | Death non-GIST cause | 3 | 33.3% |
| Mixed | 18 | 36.0% | **Median follow-up time (months)** |  |  |
| **PDGRFA mutation location** |  |  | Overall follow-up | 55.1 | - |
| Exon 12 | 5 | 10.0% | Time to metastatic disease (n=6) | 30.1 | - |
| Exon 14 | 2 | 4.0% | Metastatic disease to death (n=6) | 18.5 | - |
| Exon 18 | 43 | 86.0% |  |  |  |

**Figure 1:**
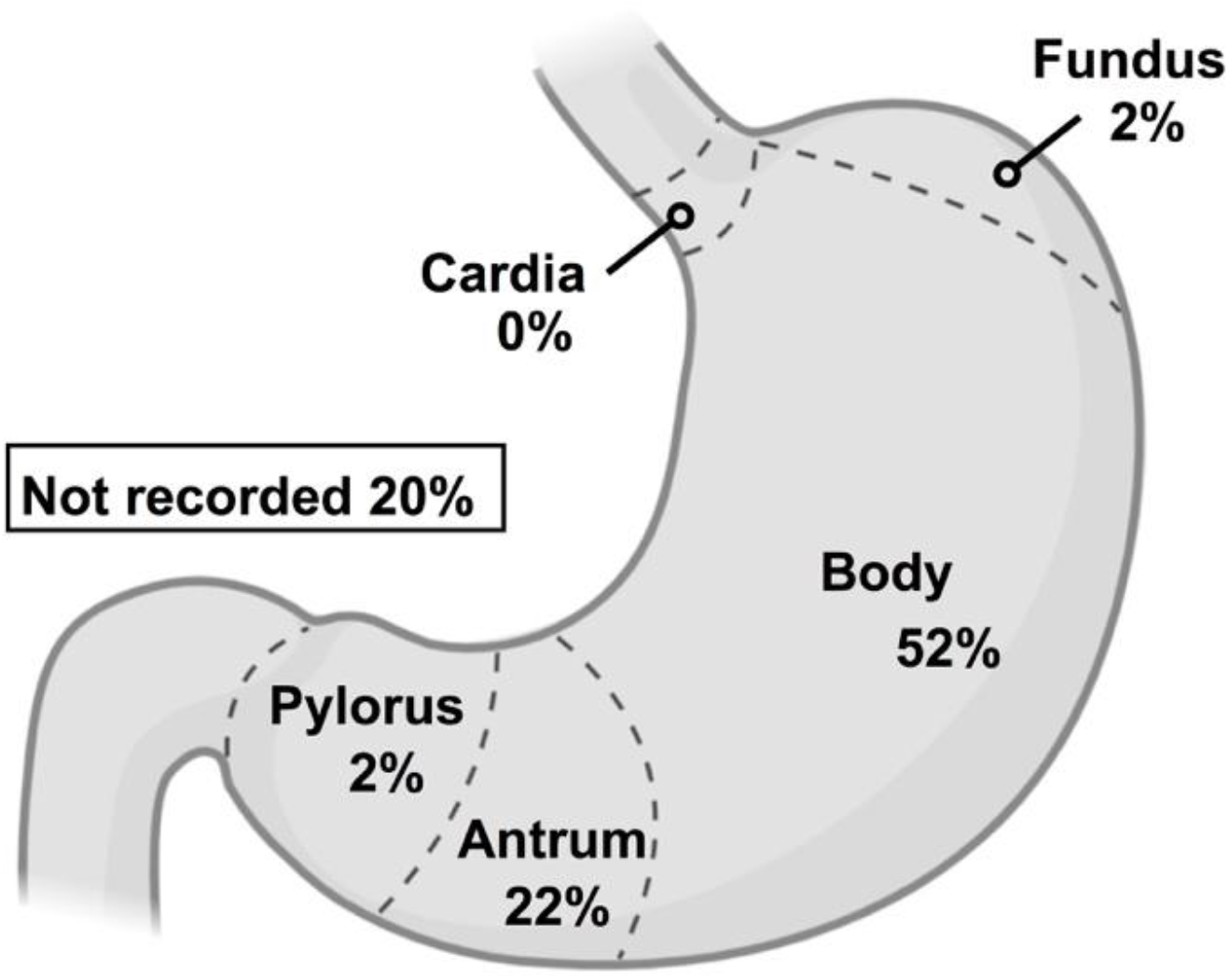
Anatomical location of all *PDGFRA*-mutant gastric GIST samples.

Most cases (46/50; 92%) were not metastatic at diagnosis. During follow-up, six patients (6/46; 13%) developed metastases with liver (8/10; 80%) and peritoneum (5/10; 50%) being the commonest sites (**Table 2**). Seven patients (7/50; 14%) received TKI therapy: one received neo-adjuvant imatinib (later receiving imatinib again for metastatic disease), two received adjuvant imatinib (3 years), and five with metastatic disease received imatinib, nilotinib, sunitinib (as part of a trial) or avapritinib (under compassionate use). Five metastatic patients did not receive any TKI therapy due to resistance prior to the availability of avapritinib.

82% of all patients remained alive at the end of the 13-year follow-up period: six patients died from metastatic GIST, whilst three died from other causes (including synchronous tumours) (**Table 2**). Median overall follow-up was 55.1 months, with the median time to metastatic disease being 30.1 months, and the median time from metastatic diagnosis to death being 18.5 months. Overall survival (OS) and time to developing metastatic disease survival curves showed a relatively indolent disease course (**Figure 2A-D, Supplementary figure 1A**) except in those who initially presented with metastatic disease (**Figure 2A**) (*p*=0.001). There was no evident difference in OS between modified AFIP stratification groups (*p*=0.5) (**Figure 2B**), time to developing metastatic disease (*p*=0.2) (**Figure 2D**), or for OS stratified by the number of mitoses (*p*=0.7) (**Supplementary Figure 1**).

**Figure 2:**
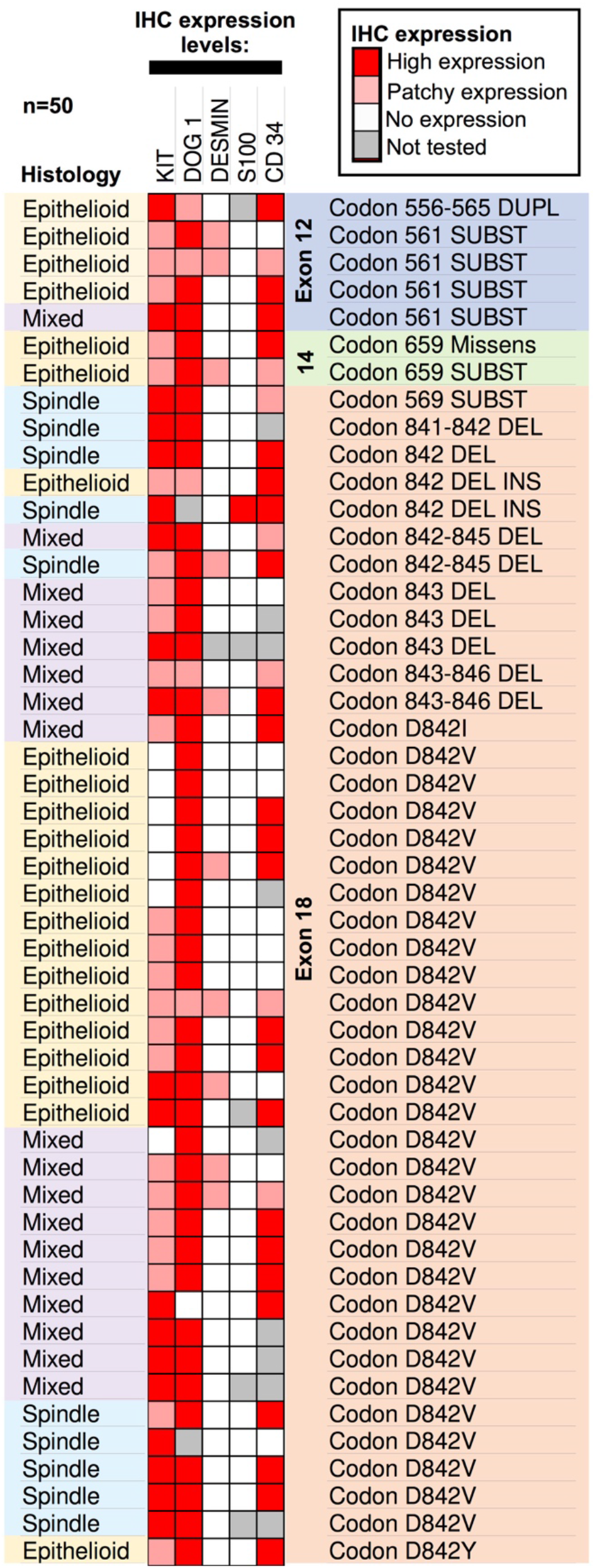
Graphical summary of all *PDGFRA*-mutant GIST samples. Each row represents an individual patient (n=50). IHC protein expression levels are displayed in the central boxes, with histological type and mutation information listed on either side (IHC: immunohistochemistry)

**Figure 3:**
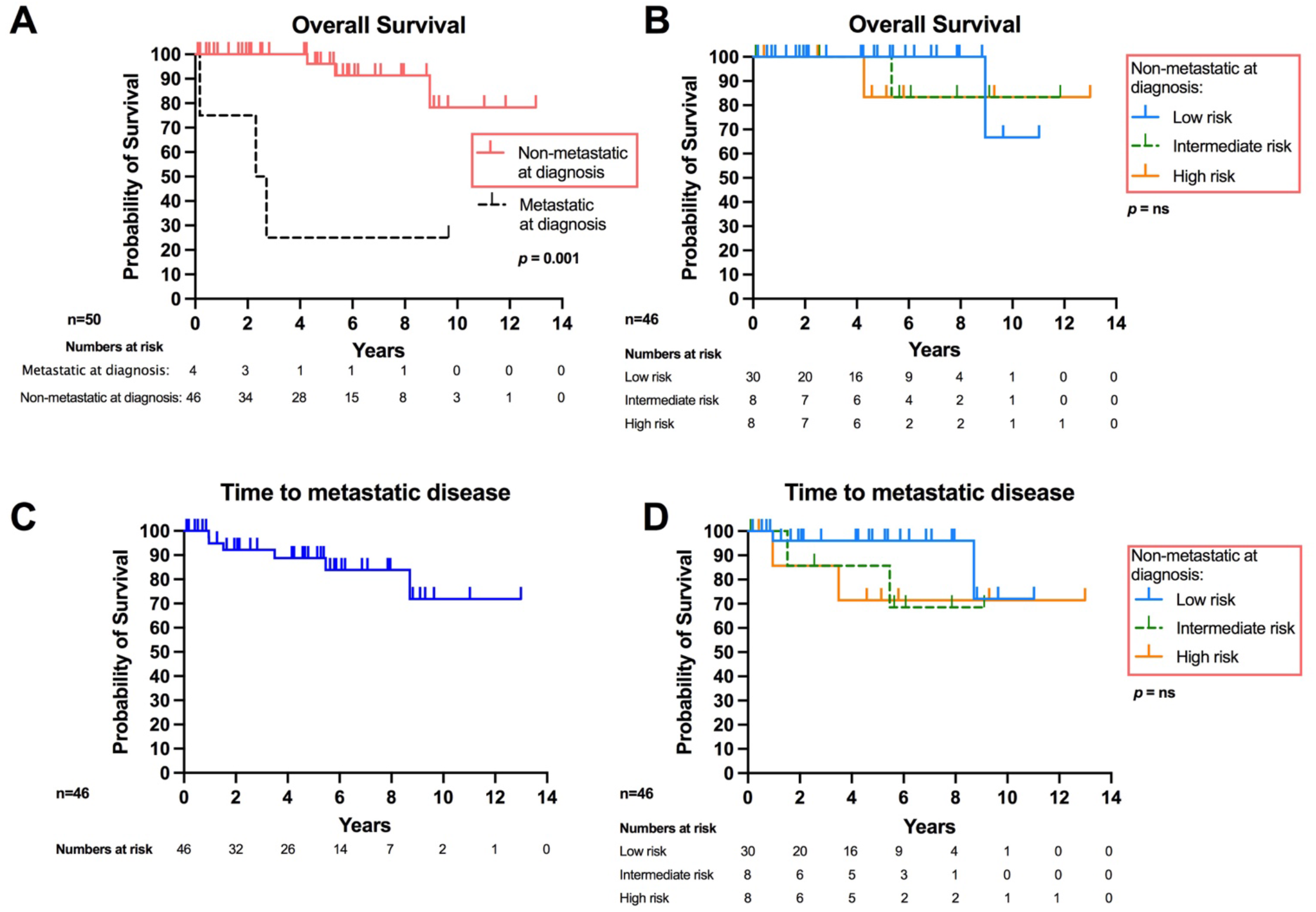
*PDGFRA*-mutant GIST survival curves. (**A)** OS for the entire cohort (n=50) stratified by presence of metastatic disease at presentation (*p*=0.001). (**B)** OS of patients who were non-metastatic at presentation (n=46), stratified by modified AFIP risk stratification (*p*=0.5). Time to developing metastatic disease for patients with non-metastatic disease at presentation **(C)** stratified by modified AFIP risk stratification grouping **(D)** (*p*=0.2). (ns=non-significant).

## Discussion

To the best of our knowledge, this 13-year retrospective *PDGRFA*-mutant GIST series (n=50) is the largest single-centre European series published to date. Key findings from this dataset include the relative indolent nature of non-metastatic *PDGFRA*-mutant GISTs, the variable levels of KIT protein expression, the majority gastric primary location as well as frequency of distribution within gastric anatomy, and male predominance.

In a recent analysis of 29 published international all-type GIST cohorts (n=13,550) the cumulative male:female distribution at diagnosis was found to be 1:1^2^. This is likely to represent the distribution for *KIT*-mutant GISTs which represent 75% of all GISTs. In *PDGFRA*-mutant GISTs however, males predominate, as shown by our study (1.5:1) and others (1.8:1)^6^. In terms of age at diagnosis and common presenting symptoms, our results (age >50 years; and GI bleed being commonest presenting symptom) fit with established trends^17 18^

In terms of anatomical location, *PDGRFA*-mutant GISTs are found almost exclusively within the stomach in contrast to *KIT*-mutant GISTs (60% gastric)^6 14^. Our study confirms this. All GISTs in our cohort presented in the stomach, majority of *PDGRFA*-mutant GISTs occur within the gastric body (72%) and antrum (28%). This adds to the findings of a recent study which found that different driver mutations influenced primary GIST location: *PDGRA*-mutant tumours (n=7) being found in the lower two-thirds whilst *KIT*-mutant tumours (n=103) were mostly located in the upper-third^19^. One patient in our cohort was diagnosed with metastatic peritoneal disease without a located primary despite extensive radiological and endoscopic investigation. Possibilities for this include that the primary was microscopic or had involuted at time of metastatic presentation.

Another interesting finding from this study was the variation in KIT expression by IHC, as previous smaller series have reported very minimal, if any, KIT positivity in *PDGFRA*-mutant GISTs^20 21^. Despite all tumours having no *KIT* mutations (assessed by mutational profiling), KIT expression levels (assessed by IHC) varied across patients with patchy expression being most common (48%), followed by high-level expression (38%) and no expression (14%). Although *KIT* and *PDGFRA* mutations are mutually exclusive in GISTs^22^, *KIT* amplification in a subgroup of *PDGFRA*-mutant GISTs may represent an alternative mechanism of tumour growth. This is suggested by a patient in our cohort (discussed below) with an imatinib-resistant *PDGFRA* exon 18 D842V mutation who also had high level KIT protein expression and benefitted from imatinib, In contrast to *PDGFRA*-mutant GISTs, *KIT*-mutant GISTS are generally KIT-positive with only 5% being KIT-negative^7^, although loss of *KIT* expression may occur following imatinib therapy as a mechanism of resistance^20^. Our DOG1 (majority positive), desmin and S100 (majority negative) IHC findings fit with established data for all GISTs, whilst CD34 expression was lower in our dataset compared to all GISTs^7 23^. Few tumours were CD34-negative and no individual tumours were both KIT and DOG1 negative.

The modified AFIP risk stratification classification (very low, low risk, intermediate risk, high risk) is commonly used to predict disease progression in treatment naive GISTs^13 14 18^. This takes into account anatomical site, tumour size and the number of mitoses as a surrogate for proliferative activity. Interestingly, AFIP risk group stratification of our study’s pre-metastatic patients did not show any statistical OS or time to metastatic disease benefit, and neither did stratification by number of mitoses alone. Although the lack of significance may be related to the small sample size, it remains unclear whether the modified AFIP stratification is as applicable to *PDGRFA*-mutant GISTs as it is in *KIT*-mutant GISTs. Meta-analysis of pooled international *PDGRFA*-mutant cohorts will be beneficial in resolving this question.

Our study’s *PDGRFA*-mutation exon frequency fits with prior studies^5^. The majority were exon 18 mutated with the commonest in this group being the imatinib-resistant D842V mutation (Figure 2). Contrasted to *KIT*-mutant GISTs, non-metastatic *PDGFRA*-mutated GISTs have a lower frequency of metastatic progression and have a generally indolent natural history^6^. This is reflected in our data as well, with the 10-year survival being in excess of 70%. A minority cohort received neoadjuvant or adjuvant TKI therapy. This was because the majority harboured imatinib-resistant mutations. In the metastatic setting, one patient with an imatinib resistant exon 18 D842V mutation received imatinib, nilotinib and sunitinib (all in the context of a clinical trial) with subsequent progression and death from metastatic disease. Another patient with aggressive metastatic disease at diagnosis commenced imatinib in a desperate attempt to contain disease before dying 1 month after starting as the results of his mutational profiling (imatinib-resistant exon 18 D842V) were only available shortly after his death (1 month after diagnosis).

Interestingly, a third patient, diagnosed with imatinib-resistant *PDGFRA-mutant* exon 18 D842V GIST had a sustained 3-year response to 400mg imatinib after developing metastatic disease. As mentioned earlier, he was one of the 38% of *PDGFRA*-mutant GISTs in our cohort with high KIT protein expression. Unfortunately, he progressed after 6 years and at time of writing was in the process of commencing avapritinib. His response to imatinib remains unexplained, possibly being related to his tumour’s high KIT expression. It also suggests that resistance may be mediated by a number of hitherto unknown factors, or that he developed metastatic disease from an alternative imatinib-sensitive GIST primary. Although there is both *in-vitro* and *in-vivo* data showing that exon 18 D842V mutations are imatinib resistant^24 25^, there is growing evidence that a small minority of D842V mutated GISTS may respond to imatinib^26^ necessitating its use in such situations in the absence of better therapeutic options.

Following its licencing by the FDA and EMA, we were able to access avapritinib on compassionate grounds for two patients with imatinib resistant exon 18 D842V mutations who developed metastatic disease and who at time of writing have had ongoing response to avapritinib for 15.1 and 7.6 months respectively. Unfortunately, despite clinical benefit in this setting^9^, the UK National Institute for Clinical Excellence (NICE) has not approved avapritinib for treatment of *PDGFRA* D842V-mutant GIST in the NHS.

### The future

The future holds much promise for *PDGFRA*-mutant GISTs, judging by the recent encouraging trial results for newer agents for avapritinib in *PDGFRA* D842V-mutant GIST and ripretinib as fourth line GIST TKI therapy. These breakthroughs suggest that a number of additional, related drugs inducing response to resistant *PDGFRA*-mutant GIST are likely to be developed as well. In order to better understand and target *PDGFRA*-mutant GISTs, future studies exploring whole genome sequencing^27^, methylation signatures^28^ and proteomics of *PDGFRA*-mutant GISTs are needed. These approaches will provide insight into the mechanisms driving these tumours and potentially open new avenues for therapeutic targeting. Similarly, there is a need for pooled meta-analyses of *PDGRFA*-mutant cohorts in order to better configure risk prediction calculators.

## Data Availability

All data produced in the present study are available upon reasonable request to the authors

## Contributors

DMF and VRB collected the data. OG performed histological and genetic analyses. DMF analysed and interpreted the data, OG and VRB provided oversight. DMF wrote the manuscript. All authors (DMF, HW, JH, OG, VRB) assisted in editing the manuscript, and all authors read and approved the final manuscript.

## Ethics approval

This study was approved by the Institutional Review Board, Addenbrookes Hospital, Cambridge University Hospitals NHS Foundation Trust, Cambridge, United Kingdom.

## Data availability statement

Data are available upon reasonable request.

## SUPPLEMENTARY DATA

**Supplementary Figure 1.**
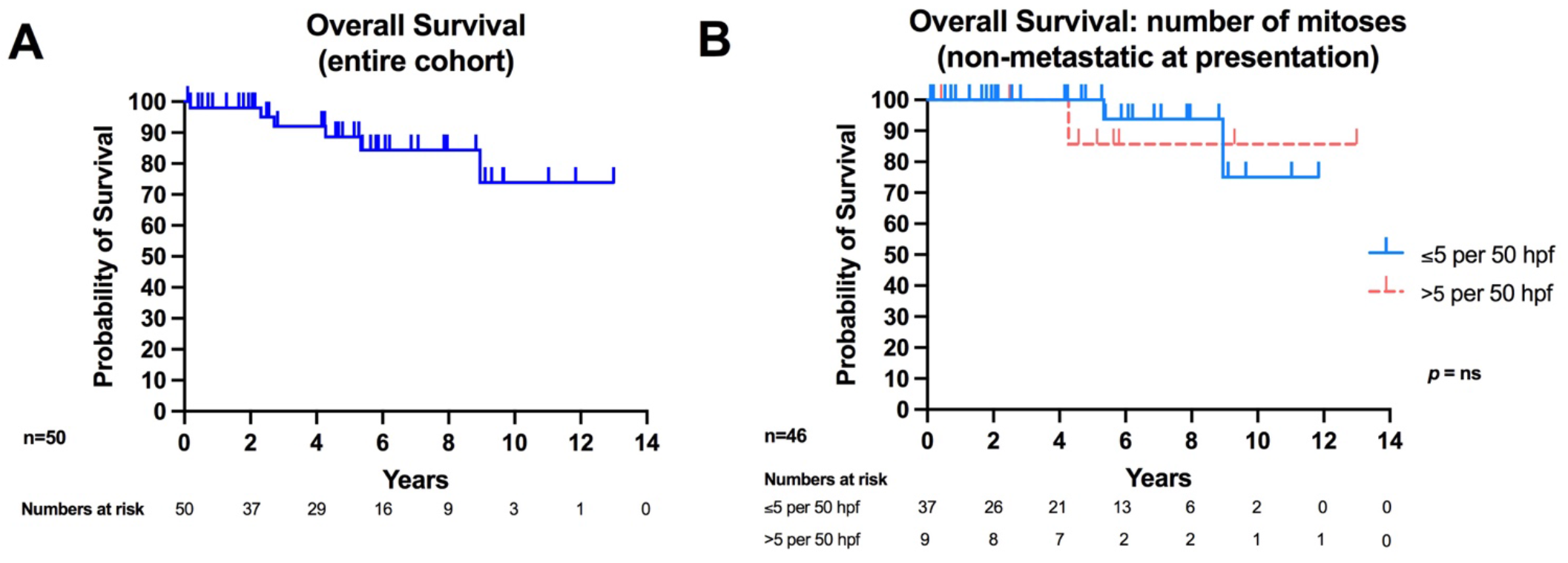
**A)** OS for the entire *PDGFRA*-mutant GIST cohort (n=50). **B)** OS for patients presenting without metastatic disease (n=46), stratified by the number of mitoses present in the primary tumour (≤ 5 per 50 hpf, versus > 5 per 50 hpf) (*p*=0.7). (Hpf=high power field; ns=non-significant)

**Supplementary Table 1.** Patient immunohistochemical (IHC) staining characteristics.

|  | n=50 | % |
| --- | --- | --- |
| **IHC: KIT (CD117)** |  |  |
| Positive | 19 | 38.0% |
| Patchy | 24 | 48.0% |
| Negative | 7 | 14.0% |
| **IHC: DOG1** |  |  |
| Positive | 42 | 84.0% |
| Patchy | 5 | 10.0% |
| Negative | 1 | 2.0% |
| Unknown | 2 | 4.0% |
| **IHC: desmin** |  |  |
| Positive | 0 | 0.0% |
| Patchy | 10 | 20.0% |
| Negative | 39 | 78.0% |
| Unknown | 1 | 2.0% |
| **IHC: S100** |  |  |
| Positive | 1 | 2.0% |
| Patchy | 0 | 0.0% |
| Negative | 44 | 88.0% |
| Unknown | 5 | 10.0% |
| **IHC: CD34** |  |  |
| Positive | 24 | 48.0% |
| Patchy | 7 | 14.0% |
| Negative | 10 | 20.0% |
| Unknown | 9 | 18.0% |

